# Exposure-Response Relation for Vibration-Induced White Finger: Inferences from a Published Meta-Analysis of Population Groups

**DOI:** 10.1101/2022.08.02.22278336

**Authors:** Magdalena F. Scholz, Anthony J. Brammer, Steffen Marburg

## Abstract

**Purpose:** It is questioned whether the exposure-response relation for the onset of vibration-induced white finger (VWF) in ISO 5349-1:2001 needs to be revised based on the epidemiologic studies identified by Nilsson et al., (2017) (https://doi.org/10.1371/journal.pone.0180795), and whether the relation they derive improves the prediction of VWF in vibrationexposed populations.

**Methods:** A pooled analysis has been performed using pidemiologic studies that complied with selection rules and reported a VWF prevalence of 10% or more, and exposure constructed according to the provisions of ISO 5349-1:2001. The lifetime exposure at 10% prevalence was calculated for various data sets using linear interpolation. They were then compared to both the model from the standard and that developed by Nilsson et al.

**Results:** Regression analyses reveal excluding extrapolation to adjust group prevalences to 10% produce models with 95-percentile confidence intervals that include the ISO exposure-response relation but not that in Nilsson et al. (2017). Different curve fits are obtained for studies involving daily exposure to single or multiple power tools and machines. Studies with similar exposure magnitudes and lifetime exposure durations but markedly different prevalences are observed to cluster.

**Conclusions:** A range of exposures and A(8)-values is predicted within which the onset of VWF is most likely to occur. The exposure-response relation in ISO 5349-1:2001, but not that proposed by Nilsson et al., falls within this range and provides a conservative estimate for the development of VWF. Additionally, the analyses demonstrate that the method for evaluating vibration exposure contained in ISO 5349-1:2001 needs revision.

## 1 Introduction

The onset of vibration-induced white finger (VWF) in workers operating power tools or machines is a subject of considerable interest for establishing occupational exposure limits. Guidelines have been proposed from epidemiologic studies and incorporated into regulations and standards. A continuing debate has focused on the accuracy of the guidelines in the international standard for hand-transmitted vibration, ISO 5349-1:2001 (ISO 5349-1, 2001), which are based on an exposure-response model developed by Brammer (1982b). In a recent comprehensive meta-analysis Nilsson et al. (2017) have analyzed data published over the last seventy years, and used a documented selection of studies to create a new model for predicting a 10% prevalence of VWF in persons whose hands are occupationally exposed to vibration. The predictions of this risk assessment model differ substantially from those contained in ISO 5349-1:2001. Here the question is raised if the model in the standard indeed needs to be revised to account for the information in the recent meta-analysis. Clearly, an accurate prediction based on an appropriate evaluation of vibration exposure is needed to protect workers from damage to the vascular, neurological and musculo-skeletal systems of their hands and to construct meaningful national regulations and legislation. Only if the effects of exposure are assessed correctly is it possible to create work environments and schedules that balance productivity with the health and safety of workers.

There have been several attempts to relate occupational exposure of the hands to vibration to the development of VWF (Bovenzi (1994), Bovenzi et al. (1995), Bovenzi (1998a), Bovenzi (2010b), Brammer (1982a), Brammer (1986), Futatsuka et al. (1984), Griffin (1982), Griffin et al. (2003) Miyashita et al. (1982), Sauni et al. (2009), Su et al. (2013), Taylor et al. (1975a), Tominaga (1982)). These range from ad hoc to population distribution driven models employing regression analyses of selected epidemiological studies of workers to logistic regression models for assessing the odds ratios associated with different methods for estimating daily and lifetime exposures. There are also models for longitudinal studies.

It is evident from this body of work that establishing a relation between vibration exposure and the development of vascular or neurological disturbances in the hand encounters several difficulties. These relate to the measurement of vibration at the hands, determining the exposure to it, ergonomic factors such as hand force and grip, and posture, and relying on information given by participants concerning their signs, symptoms and work history. Furthermore, different measures of exposure, such as the lifetime vibration dose (Griffin et al., 2003), cumulative exposure index (Sauni et al., 2009) or total operating time (Miyashita et al., 1982) provide alternate and not always compatible metrics for evaluating or predicting the harm from vibration exposure. Additionally, most exposures have been in a temperate climate and there are relatively few in a tropical climate. It is well known that low temperatures can cause fingers to whiten and hence vascular spasms are more likely to occur in hands affected by VWF than when in a near-tropical climate (Futatsuka et al. (2005), Su et al. (2013)).

An analysis of the relative weight to apply to the magnitude of vibration at a surface in contact with the hands compared to the lifetime exposure duration found that a better prediction of the health effects could be obtained by applying the same power to the total exposure time as to the vibration magnitude, in contrast to the method contained in ISO 5349-1:2001 (Griffin et al., 2003). The authors related this to the calculation not distinguishing between exposures accumulated over a day and those over several years. Also, Griffin et al. found that using an unweighted acceleration resulted in a better prediction of VWF than if the frequency-weighted acceleration recommended in the ISO standard was used. However, recent work has shown that a more nuanced approach is needed to specify a frequency-weighting for at least the vascular component of hand-arm vibration syndrome (HAVS) (Brammer and Pitts, 2012). Studies have also reported that the relation between vibration exposure and the development of VWF contained in the international standard both underestimates or overestimates the risk in different population groups (Bovenzi et al. (1988), Bovenzi et al. (1995), Bovenzi (1998a), Bovenzi (2012), Engström and Dandanell (1986), Futatsuka et al. (1984),Gerhardsson et al. (2020), Keith and Brammer (1994), Starck et al. (1990), Tominaga (1990), Walker et al. (1985)).

In light of these findings and the availability, for the first time, of a comprehensive, systematic meta-analysis identifying studies conducted over the last seventy years relating occupational exposure to vibration to the development of VWF, it would appear both imperative and timely to reassess the suitability of the international standard for the purpose for which it was designed. Accordingly, the purpose of this contribution may be summarized in two objectives. The first is to examine whether the exposure-response relation for the onset of VWF contained in ISO 5349-1:2001, including the method for calculating exposure, needs to be revised based on the results of epidemiologic studies included in the recent meta-analysis by Nilsson et al. (2017). The second is to consider whether the exposure-response relation proposed by Nilsson et al. (2017) improves the prediction of VWF in vibration-exposed population groups. Answers to these questions could imply a need to revise not only the model contained in the standard, but also of the methods for evaluating exposure. Such revisions would influence implementation of regulations limiting workplace vibration exposure and machinery vibration emission in many countries that are dependent on the standard, with immediate health and economic consequences.

In this study, the approach chosen by Nilsson et al. (2017) is replicated with modifications to create models to predict the prevalence of 10% white fingers in a population group for a given vibration exposure, as described in the Method section. The models are constructed using the procedures for estimating daily and lifetime exposures contained in the international standard. They are then described in the Results with both the exposure-response relation from the standard and that developed by Nilsson et al. (2017). The relation of the three models to the epidemiologic data is analyzed in the Discussion, together with the limitations of the study, to address whether the model in the ISO 5349-1:2001 needs revision and if the model created by Nilsson et al. (2017) is an improvement on that in the standard.

## 2 Method

The meta-analysis performed by Nilsson et al. (2017) consisted of a systematic review of original scientific papers published in English in refereed journals. Screening of the literature was initially done by abstract followed by an evaluation of 294 articles against pre-determined criteria established to evaluate overall “quality”, from which 52 were judged to be of sufficient quality for inclusion in the analysis. The criteria involved are described in detail in the Annex of Nilsson et al. (2017). Out of these 41 contained data regarding Raynaud’s phenomenon.

An overview of the data from all publications judged acceptable by Nilsson et al. (2017) was first created, including the methods of exposure measurement and clinical evaluation used in the various studies. These data were then further screened by the selection rules introduced here (table 1) to reduce heterogeneity and so ensure compatibility with the objectives of the present study. Hence for the purposes of the present pooled analyses, hand-transmitted vibration had to have been measured in accordance with the requirements of the international standard in effect at the time of the study (ISO 5349:1986, or ISO 5349-1:2001). This is ensured if the first five conditions in table 1 are fulfilled. Regarding epidemiologic data, studies of VWF are to be included in the analyses if they satisfy conditions 6 to 11. These rules are designed to ensure that groups are comparable and only those are included in which white fingers are caused by vibration. For example, rule 7 states that the first episode of finger blanching should occur at a fingertip after commencing occupational exposure to hand-transmitted vibration, which is a typical characteristic that distinguishes VWF from white fingers caused by unrelated disease or other factors (Taylor et al., 1975b). Furthermore, as argued in Brammer (1982b), the population group size has to be considered when evaluating such data, as small groups may not be representative of a larger population. It was found there that consistency in the data analysis was obtained for a minimum group size of thirty persons, which is included here in rule 6. The selection rules are intended to enable a simple binary decision between whether or not to include the results of a study in the analyses. However, there were a few studies that may or may not comply with all selection rules on which judgments had to be made concerning the reliability of the data.

**Table 1.**
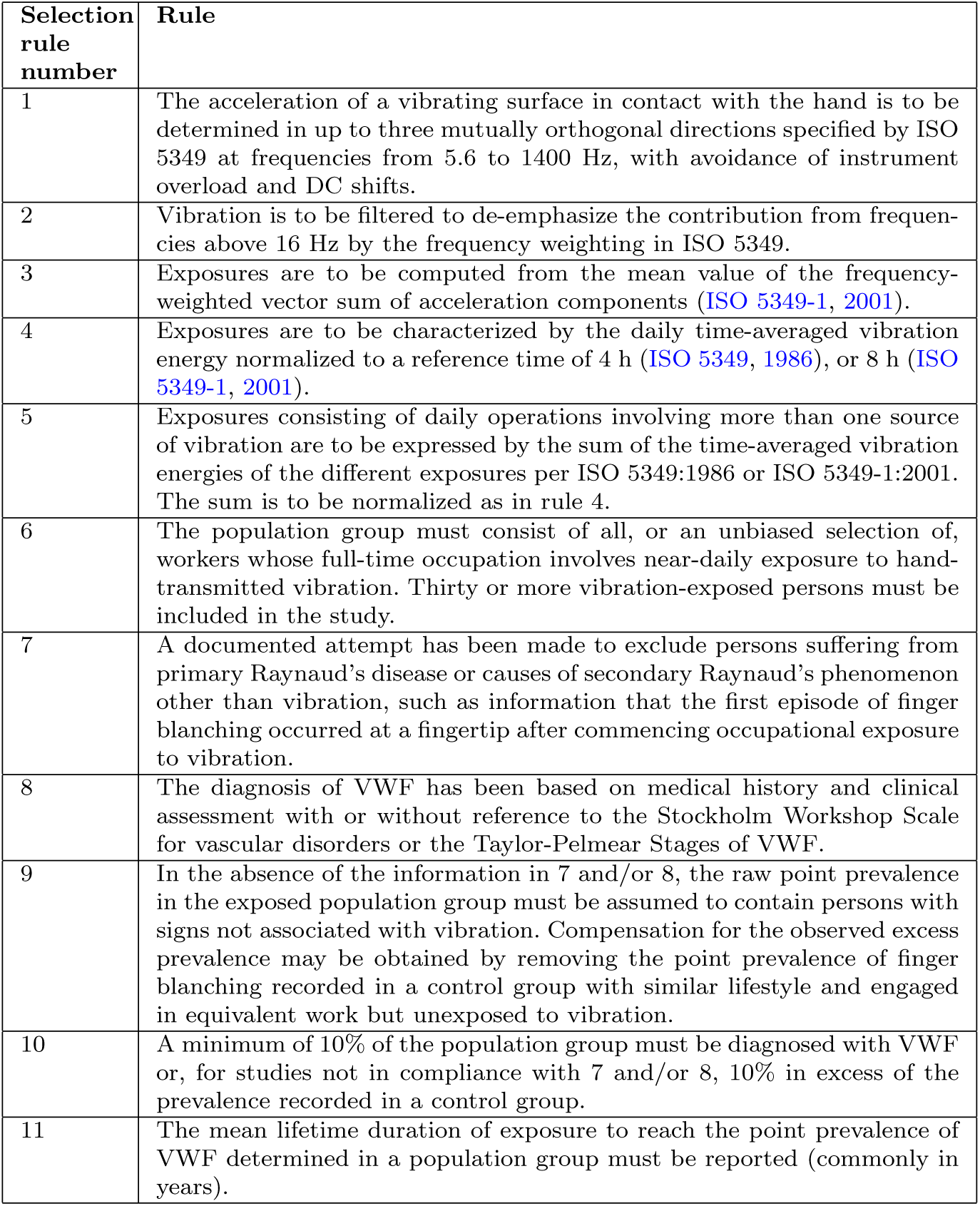
Selection rules used to determine the reliability of the data provided by the studies and hence their usage in this pooled analysis (Rules 1-5: measurement of vibration, 6-11: epidemiologic data)

| Selection rule number | Rule |
| --- | --- |
| 1 | The acceleration of a vibrating surface in contact with the hand is to be determined in up to three mutually orthogonal directions specified by ISO 5349 at frequencies from 5.6 to 1400 Hz, with avoidance of instrument overload and DC shifts. |
| 2 | Vibration is to be filtered to de-emphasize the contribution from frequencies above 16 Hz by the frequency weighting in ISO 5349. |
| 3 | Exposures are to be computed from the mean value of the frequency-weighted vector sum of acceleration components (ISO 5349-1, 2001). |
| 4 | Exposures are to be characterized by the daily time-averaged vibration energy normalized to a reference time of 4 h (ISO 5349, 1986), or 8 h (ISO 5349-1, 2001). |
| 5 | Exposures consisting of daily operations involving more than one source of vibration are to be expressed by the sum of the time-averaged vibration energies of the different exposures per ISO 5349:1986 or ISO 5349-1:2001. The sum is to be normalized as in rule 4. |
| 6 | The population group must consist of all, or an unbiased selection of, workers whose full-time occupation involves near-daily exposure to hand-transmitted vibration. Thirty or more vibration-exposed persons must be included in the study. |
| 7 | A documented attempt has been made to exclude persons suffering from primary Raynaud's disease or causes of secondary Raynaud's phenomenon other than vibration, such as information that the first episode of finger blanching occurred at a fingertip after commencing occupational exposure to vibration. |
| 8 | The diagnosis of VWF has been based on medical history and clinical assessment with or without reference to the Stockholm Workshop Scale for vascular disorders or the Taylor-Pelmeur Stages of VWF. |
| 9 | In the absence of the information in 7 and/or 8, the raw point prevalence in the exposed population group must be assumed to contain persons with signs not associated with vibration. Compensation for the observed excess prevalence may be obtained by removing the point prevalence of finger blanching recorded in a control group with similar lifestyle and engaged in equivalent work but unexposed to vibration. |
| 10 | A minimum of 10% of the population group must be diagnosed with VWF or, for studies not in compliance with 7 and/or 8, 10% in excess of the prevalence recorded in a control group. |
| 11 | The mean lifetime duration of exposure to reach the point prevalence of VWF determined in a population group must be reported (commonly in years). |

The exposure-response relation in ISO 5349-1:2001 predicts the mean time exposed (in years) to a daily exposure characterized by the 8-h energy-equivalent averaged acceleration, A(8), for the prevalence of VWF in a population group to reach 10%, where:

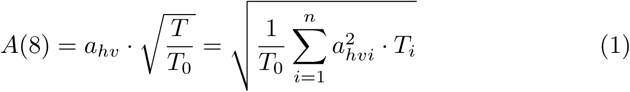

In this equation *T*_0_ is the reference time for calculating the daily exposure, which is 8 hours in ISO 5349-1:2001 in order to represent a conventional workday. The earlier version of the standard employed a reference time of 4 hours, but this is not applied to data here. *T* is the time the users were exposed to the frequency-weighted vibration total value, *a*_*hv*_. For daily exposures involving a variety of power tools or machines, each used for different times during a workday, the component exposures are summed as in equation 1, where *T*_*i*_, is the time of exposure to the i^*th*^ tool or machine with vibration total value of *a*_*hvi*_. Now the point prevalence of VWF recorded in all epidemiologic studies included in the analyses was not 10%. Thus, in order to examine the accuracy of the ISO prediction, the exposure duration at the point in time of each study at which 10% of the population would have been affected by VWF needs to be determined. For this purpose, the method used in Nilsson et al. (2017), which assumes a linear increase in prevalence of Raynaud’s phenomenon with time, is also used here. But in order to keep the error as low as possible, extrapolation is avoided and only interpolation allowed. Consequently, only data for those population groups that had a prevalence of 10% or more when the epidemiologic study was conducted are included in the analyses.

Furthermore, a zero prevalence of VWF at zero duration lifetime exposure needs to be assumed in order to reconstruct the lifetime exposure for 10% prevalence. This implies that the observed prevalence of VWF contains no individuals with signs and symptoms from causes other than vibration exposure, which is the reason for selection rules 7, 9 and 10 (table 1). Rule 7 requires a differential diagnosis to rule out other causes for white finger for cases observed in a given study. If there is doubt surrounding the origin of the white fingers reported, rule 9 requires an unexposed control group to be a part of each study to enable the raw prevalence to be adjusted. An adjustment is only made in the analyses described here if the authors were not convinced that the conditions contained in the rules had been met.

Most of the studies used in Nilsson et al. (2017) involved population groups that operated more than one vibrating power tool or machine per workday: hence exposures to multiple tools are included as long as rule 5 is satisfied, with A(8) calculated according to equation 1.

In addition to estimating the exposure time at 10% point prevalence, in some cases more calculation was needed in order to have data in the format needed for the models. Chatterjee et al. (1978) did not provide an A(8)-value, but published vibration spectra from which it could be calculated. Numerical values were recovered from the spectra in the graphs showing the measured vibration in (Chatterjee et al., 1978). Using these a frequency weighted spectrum was determined. And the *a*_*hv*_-value was calculated by forming the vector sum of the acceleration components according to equation 2 (from ISO 5349-1:2001) and inserting it into equation 1.

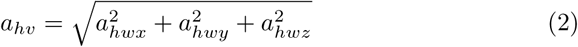

The other study for which additional calculations were needed to reduce heterogeneity is that by Bovenzi (1998b). Here, the lifetime exposure is given in total hours of tool or machine usage. Thus, it needs to be converted into the corresponding exposure in years in order to be usable in the models. Therefore, the number of workdays was estimated from statistics for the average number of workdays per year from 1965 to 1994 in the country concerned (The Workingdays Team, 15.01.2021). Twenty statutory vacation days as well as the average number of sick days derived from WHO statistics were deducted from this number to estimate the average number of days actually worked annually (World Health Organization, 15.01.2021). These workdays were then used to calculate the lifetime exposure in years:

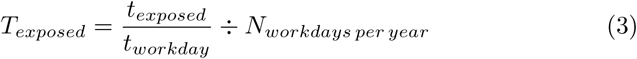

In this equation *T*_*exposed*_ is the time the workers were exposed given in years, *t*_*exposed*_ is the total hours of tool or machine usage and *t*_*workday*_ equals 8 hours. The division of *t*_*exposed*_ by *t*_*workday*_ gives the total number of workdays the users were exposed. Dividing this by the average number of days worked per year, *N*_*workdays per year*_, enables *T*_*exposed*_ to be estimated.

These calculations, together with linear interpolation of the point prevalence, produced a data set of mean group lifetime exposures in years to reach 10% prevalence for the corresponding A(8)-values. This data set was then analyzed using a regression analysis. Hence, the following power function was used, which also has the same form as the model employed in the standard ISO 5349-1 (2001):

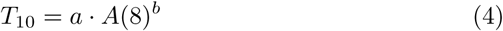

where *T*_10_ is the mean cumulative lifetime exposure of the population group to reach a 10% prevalence of VWF, and *a* and *b* are best-fit numerical parameters to the data. This process was done first with the data subset for which no calculations except interpolation to 10% prevalence were needed. Then the subsets that required further calculations were added stepwise, starting with Chatterjee et al. (1978) and then Bovenzi (1998b). Additionally, studies using only a single power tool or machine were analyzed separately. In all cases, no limitation on daily exposure duration was set.

The objectives of this study are achieved by interpreting the results of the regression analyses. Particular attention is paid to the gradient of the relations, *b*, and the goodness of fit of the models to the data (coefficient of determination, *r*^2^). Additionally, 95-percentile confidence intervals for the functional relations are presented, i.e., the intervals delineate the range of relations with form given by equation 4 that are compatible with the data. The confidence intervals thus provide tests for the accuracy of the model included in the international standard and provide information on whether the exposure-response relation proposed by Nilsson et al. (2017) improves prediction of the development of VWF in vibration-exposed population groups.

## 3 Results

Studies deemed reliable by Nilsson et al. (2017) for evaluating the development of VWF and available to the present authors are listed in table 2. The reason for exclusion from the analyses is given, and whether an A(8)-value was provided for the group’s exposure. The last two columns indicate whether and what further processing of the data was required, and if an adjustment to the reported prevalence of VWF in the population group was needed.

**Table 2.** Studies considered usable by Nilsson et al. (2017) with the reason if used or not in the analyses, whether an A(8)-value is provided, whether further processing of the data is needed to get the A(8)-value or the exposure time needed for the model

| Study | In-<br>cluded | Reason for in-<br>/exclusion | A(8)<br>pro-<br>vided | Further<br>Process-<br>ing &<br>Method |
| --- | --- | --- | --- | --- |
| Bovenzi et al. (1980) | no | no acceleration<br>value(s) | no | - |
| Bovenzi et al. (1985) | no | no mechanical filter | no | no |
| Bovenzi et al. (1988) | no | no A(8) | no | no |
| Bovenzi (1994) | yes | - | yes | no |
| Bovenzi et al. (1995) | yes | - | yes | no |
| Bovenzi (1998b) | yes | - | yes | yes, see eq.<br>3 |
| Bovenzi et al. (2005) | no | less than 10% preva-<br>lence | yes | no |
| Bovenzi (2008) | yes | - | yes | no |
| Bovenzi et al. (2008) | yes | - | yes | no |
| Bovenzi (2010a) | yes | - | yes | no |
| Bovenzi (2010b) | no | same data as<br>Bovenzi (2010a) | yes | - |
| Burström et al. (2010) | no | no acceleration<br>value(s) | no | - |
| Hagberg et al. (2008) | no | missing data | no | - |
| Nilsson et al. (1989) | no | no A(8) | no | no |
| Brubaker et al. (1987) | no | no control group | no | - |
| Chatterjee et al. (1978) | yes | vibration spectra<br>show no evidence<br>of distortion due<br>to lack of mechan-<br>ical filter during<br>measurement | no | yes, see eq.<br>1 & 2 |
| Letz et al. (1992) | no | no acceleration<br>value(s) | no | no |
| Palmer et al. (1998) | no | no control group | no | - |
| Walker et al. (1985) | no | no vibration<br>value(s) | no | - |
| Mirbod et al. (1992) | no | no unexposed con-<br>trol group | no | - |
| Mirbod et al. (1994) | no | vibration measure-<br>ment method | no | - |
| Yamada et al. (1995) | no | no control group | no | - |
| Tominaga (1994) | no | no control group | no | - |
| Su et al. (2013) | no | no control group | no | - |

The decision on inclusion or exclusion of a study was based on the selection rules in table 1. Yet, as stated in the method section, not all studies fit into this binary framework. For example, according to the description of the measurement procedure in Chatterjee et al. (1978), no mechanical filter was used despite measuring the vibration of percussive tools. This omission would exclude the study according to the selection rules. However, the authors provide vibration spectra for the power tools from which it can be seen there is no spurious increase in acceleration with decreasing frequency (i.e., no evidence of “DC-Shift”), and hence no evidence of perturbed acceleration values. For these reasons the study is included in the analyses.

The data from the studies marked as included in table 2 are listed in table 3 and plotted in figures 1 to 4. Figures 1 to 3 show the mean lifetime exposure to reach 10% point prevalence of white fingers in a vibration-exposed population group as a function of the daily exposure expressed by A(8). The model from ISO 5349-1:2001 is included in all the figures as a dashed black line. The model created by Nilsson et al. (2017) is plotted as a continuous red line. The best fit to the data is shown by the continuous blue line, and the thick blue lines display the 95-percentile confidence intervals for the regression.

**Table 3.**
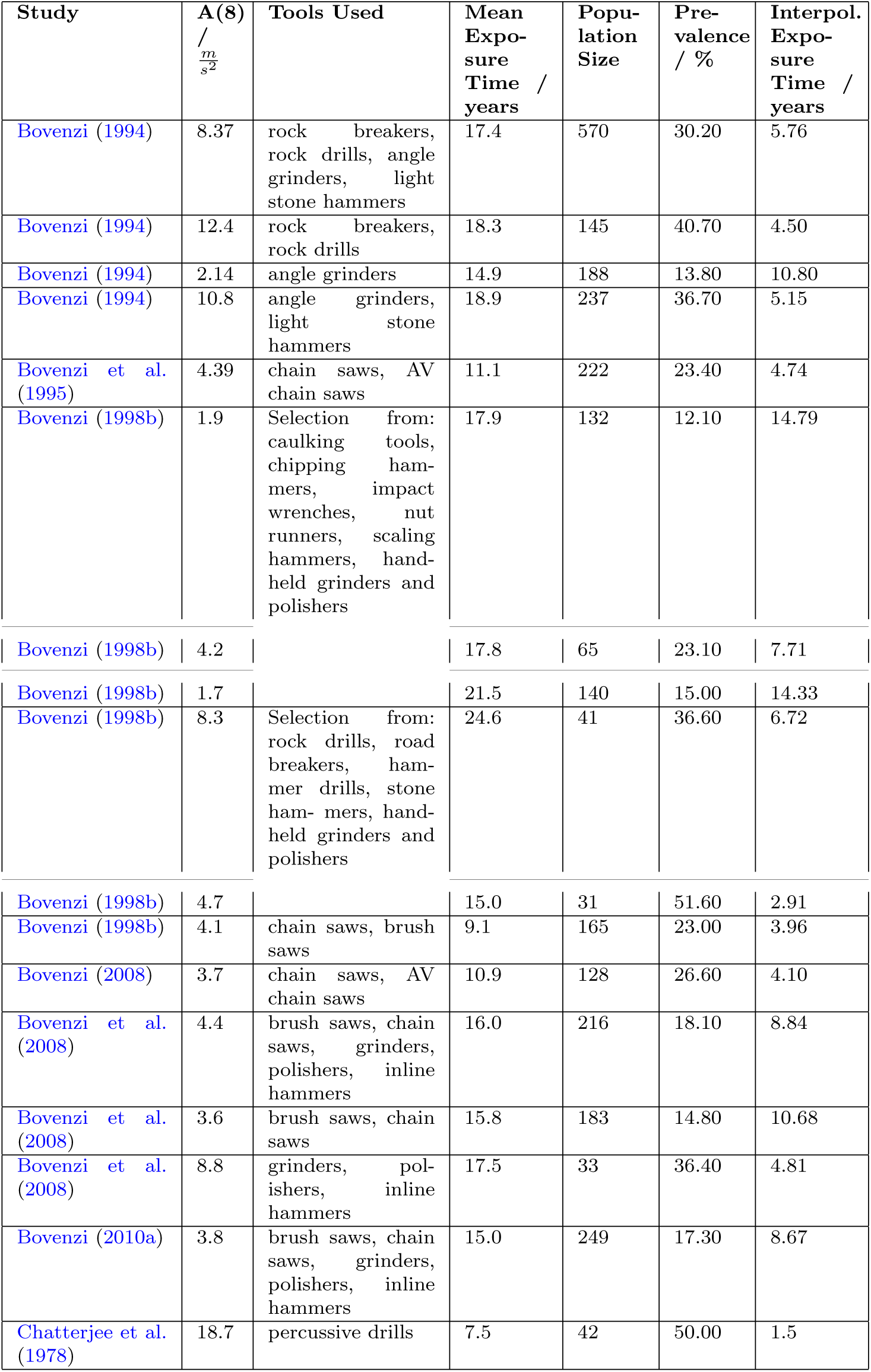
Tools used, A(8)-values, mean group exposure times, population size and point prevalences derived from the publications, and the interpolated exposure times to 10% point prevalence. All data for male workers.

| Study | A(8)<br>/<br>$\frac{m}{s^2}$ | Tools Used | Mean<br>Expo-<br>sure<br>Time /<br>years | Popu-<br>lation<br>Size | Pre-<br>valence<br>/ % | Interpol.<br>Expo-<br>sure<br>Time /<br>years |
| --- | --- | --- | --- | --- | --- | --- |
| Bovenzi (1994) | 8.37 | rock breakers,<br>rock drills, angle<br>grinders, light<br>stone hammers | 17.4 | 570 | 30.20 | 5.76 |
| Bovenzi (1994) | 12.4 | rock breakers,<br>rock drills | 18.3 | 145 | 40.70 | 4.50 |
| Bovenzi (1994) | 2.14 | angle grinders | 14.9 | 188 | 13.80 | 10.80 |
| Bovenzi (1994) | 10.8 | angle grinders,<br>light stone<br>hammers | 18.9 | 237 | 36.70 | 5.15 |
| Bovenzi et al.<br>(1995) | 4.39 | chain saws, AV<br>chain saws | 11.1 | 222 | 23.40 | 4.74 |
| Bovenzi (1998b) | 1.9 | Selection from:<br>caulking tools,<br>chipping ham-<br>mers, impact<br>wrenches, nut<br>runners, scaling<br>hammers, hand-held grinders and<br>polishers | 17.9 | 132 | 12.10 | 14.79 |
| Bovenzi (1998b) | 4.2 |  | 17.8 | 65 | 23.10 | 7.71 |
| Bovenzi (1998b) | 1.7 |  | 21.5 | 140 | 15.00 | 14.33 |
| Bovenzi (1998b) | 8.3 | Selection from:<br>rock drills, road<br>breakers, ham-<br>mer drills, stone<br>ham- mers, hand-<br>held grinders and<br>polishers | 24.6 | 41 | 36.60 | 6.72 |
| Bovenzi (1998b) | 4.7 |  | 15.0 | 31 | 51.60 | 2.91 |
| Bovenzi (1998b) | 4.1 | chain saws, brush<br>saws | 9.1 | 165 | 23.00 | 3.96 |
| Bovenzi (2008) | 3.7 | chain saws, AV<br>chain saws | 10.9 | 128 | 26.60 | 4.10 |
| Bovenzi et al.<br>(2008) | 4.4 | brush saws, chain<br>saws, grinders,<br>polishers, inline<br>hammers | 16.0 | 216 | 18.10 | 8.84 |
| Bovenzi et al.<br>(2008) | 3.6 | brush saws, chain<br>saws | 15.8 | 183 | 14.80 | 10.68 |
| Bovenzi et al.<br>(2008) | 8.8 | grinders, pol-<br>ishers, inline<br>hammers | 17.5 | 33 | 36.40 | 4.81 |
| Bovenzi (2010a) | 3.8 | brush saws, chain<br>saws, grinders,<br>polishers, inline<br>hammers | 15.0 | 249 | 17.30 | 8.67 |
| Chatterjee et al.<br>(1978) | 18.7 | percussive drills | 7.5 | 42 | 50.00 | 1.5 |

**Fig. 1.**
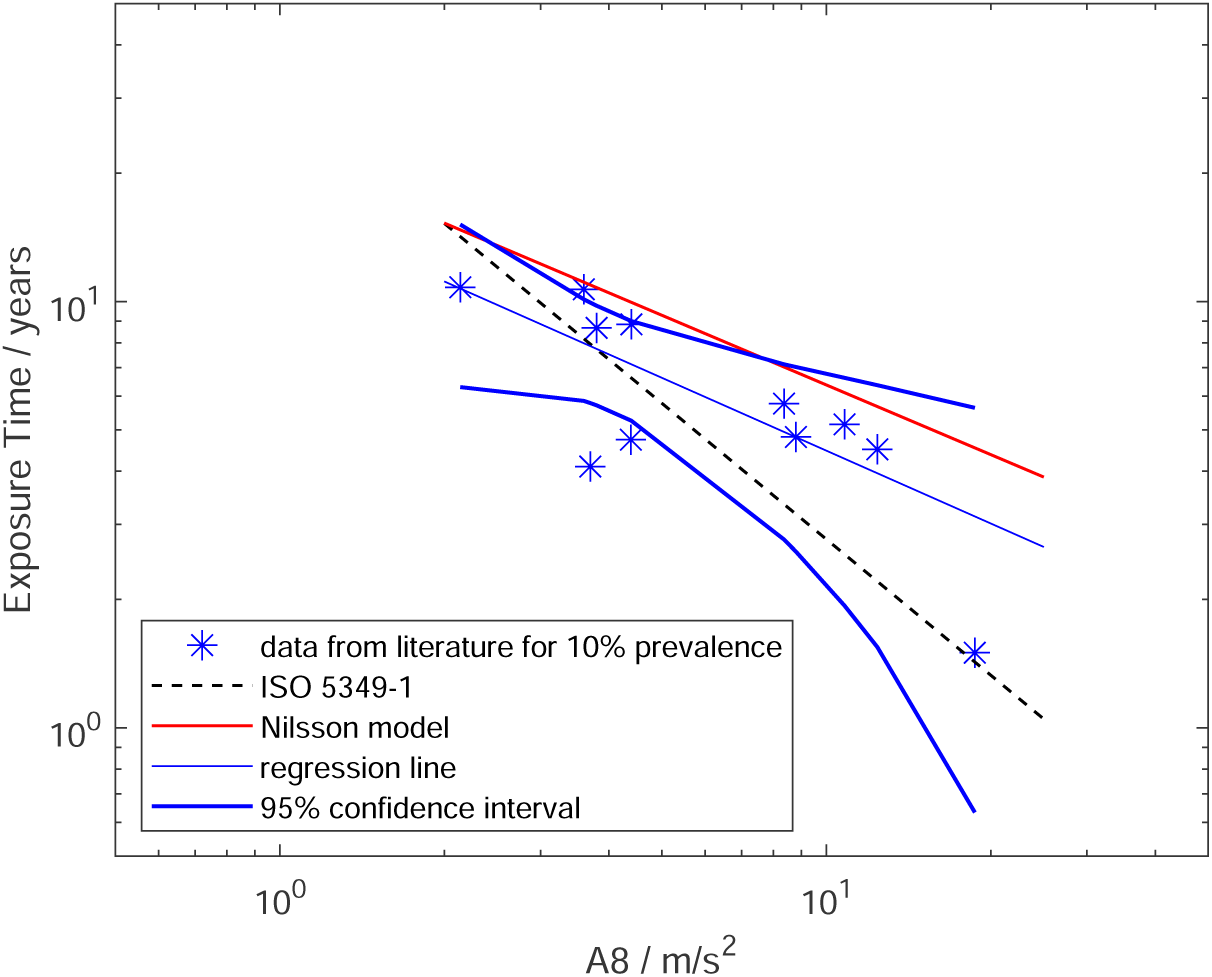
Model 1: Predicted mean lifetime exposures versus A(8) to reach 10% point prevalence of VWF for data set (stars), regression line for model 1 (continuous blue line), model from Nilsson et al. (2017) (red line), exposure-response relation from ISO 5349-1:2001 (dashed black line), and 95-percentile confidence intervals for model 1 (thick blue lines)

**Fig. 2.**
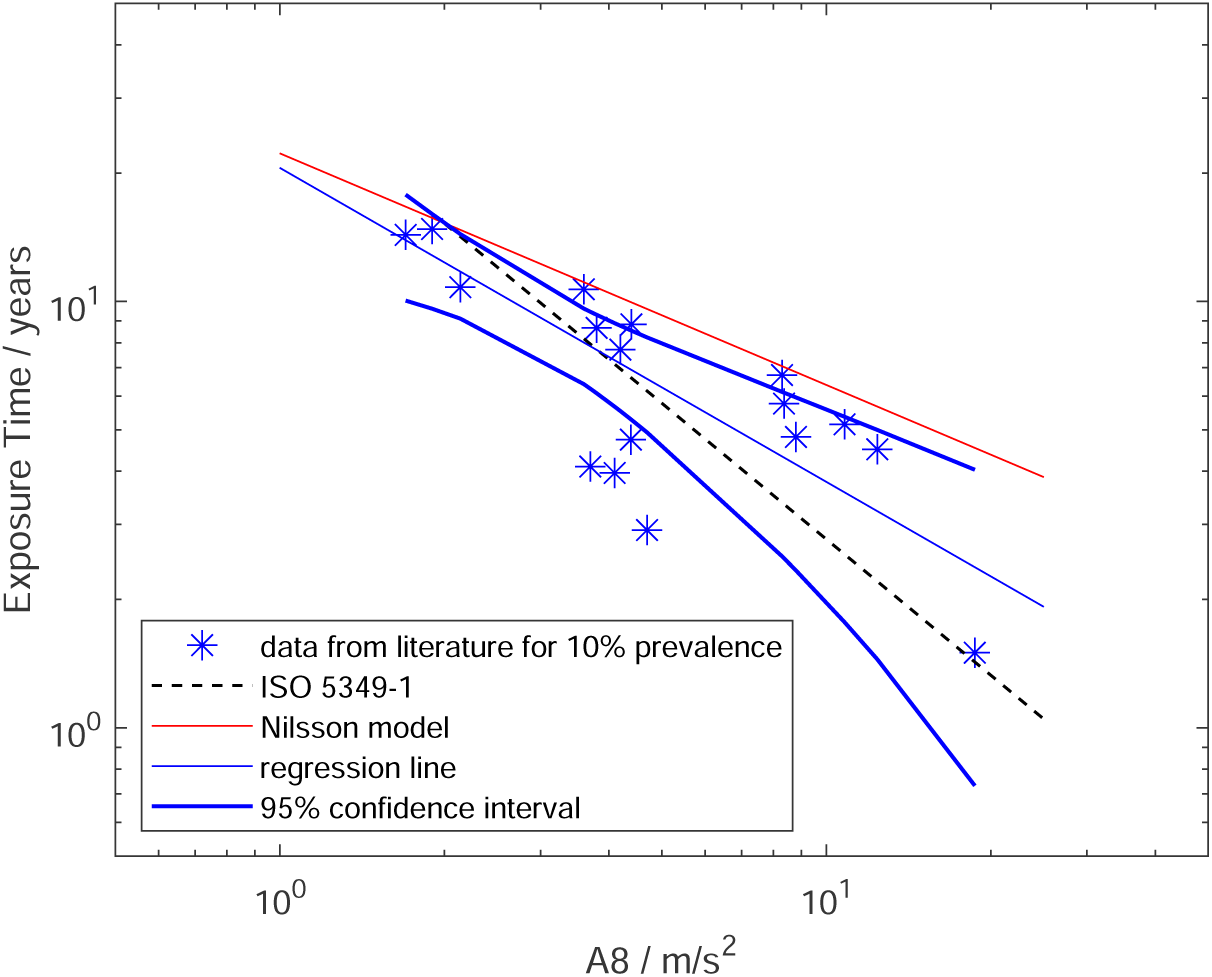
Model 2: Predicted mean lifetime exposures versus A(8) to reach 10% point prevalence of VWF for data set (stars), regression line for model 2 (continuous blue line), model from Nilsson et al. (2017) (red line), exposure-response relation from ISO 5349-1:2001 (dashed black line), and 95-percentile confidence intervals for model 2 (thick blue lines)

**Fig. 3.**
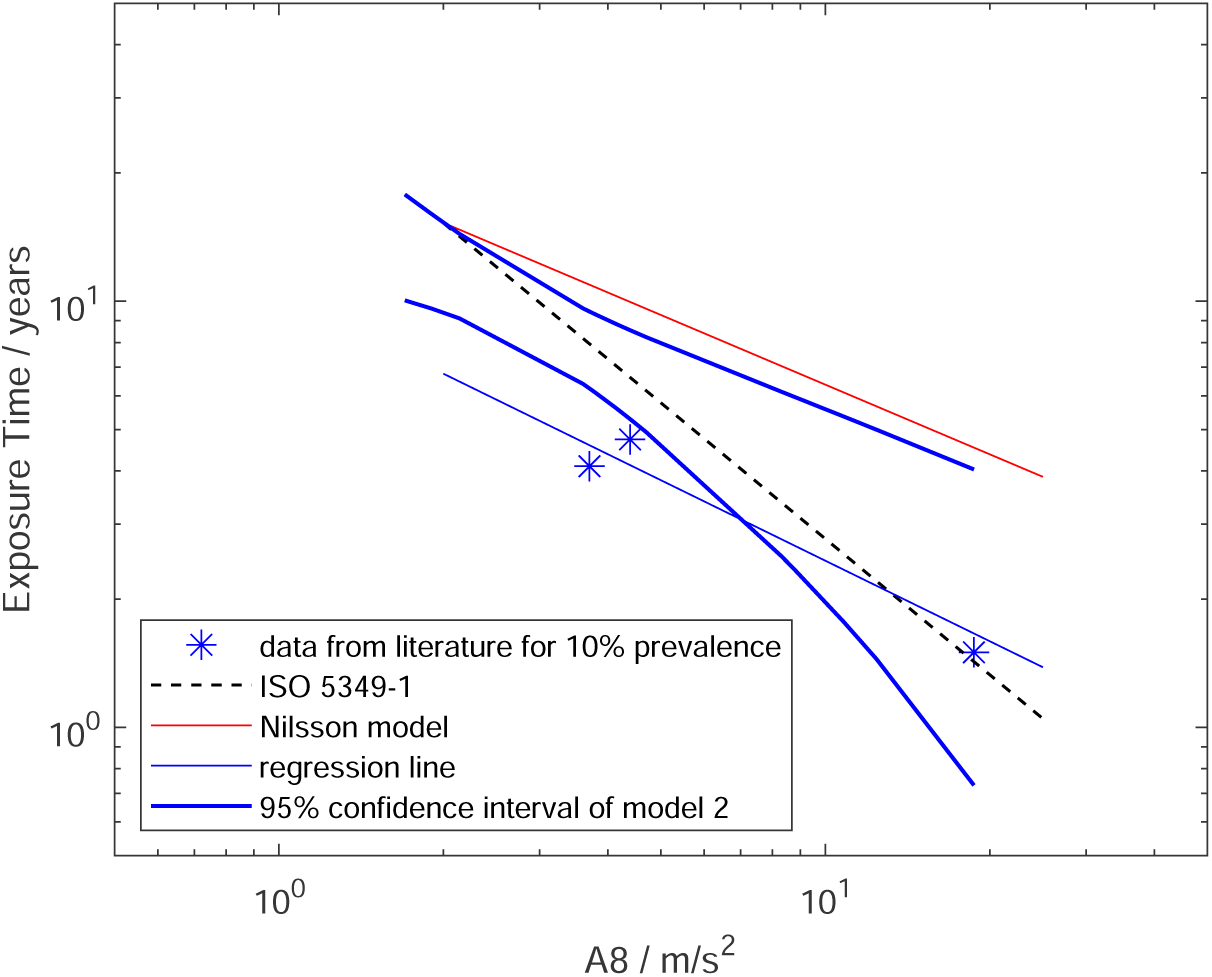
Model 3: Predicted mean lifetime exposures versus A(8) to reach 10% point prevalence of VWF for studies in which workers only used one power tool throughout the workday (stars), regression line for model 3 (continuous blue line), model from Nilsson et al. (2017) (red line), exposure-response relation from ISO 5349-1:2001 (dashed black line), and 95-percentile confidence intervals for model 2 (thick blue lines)

**Fig. 4.**
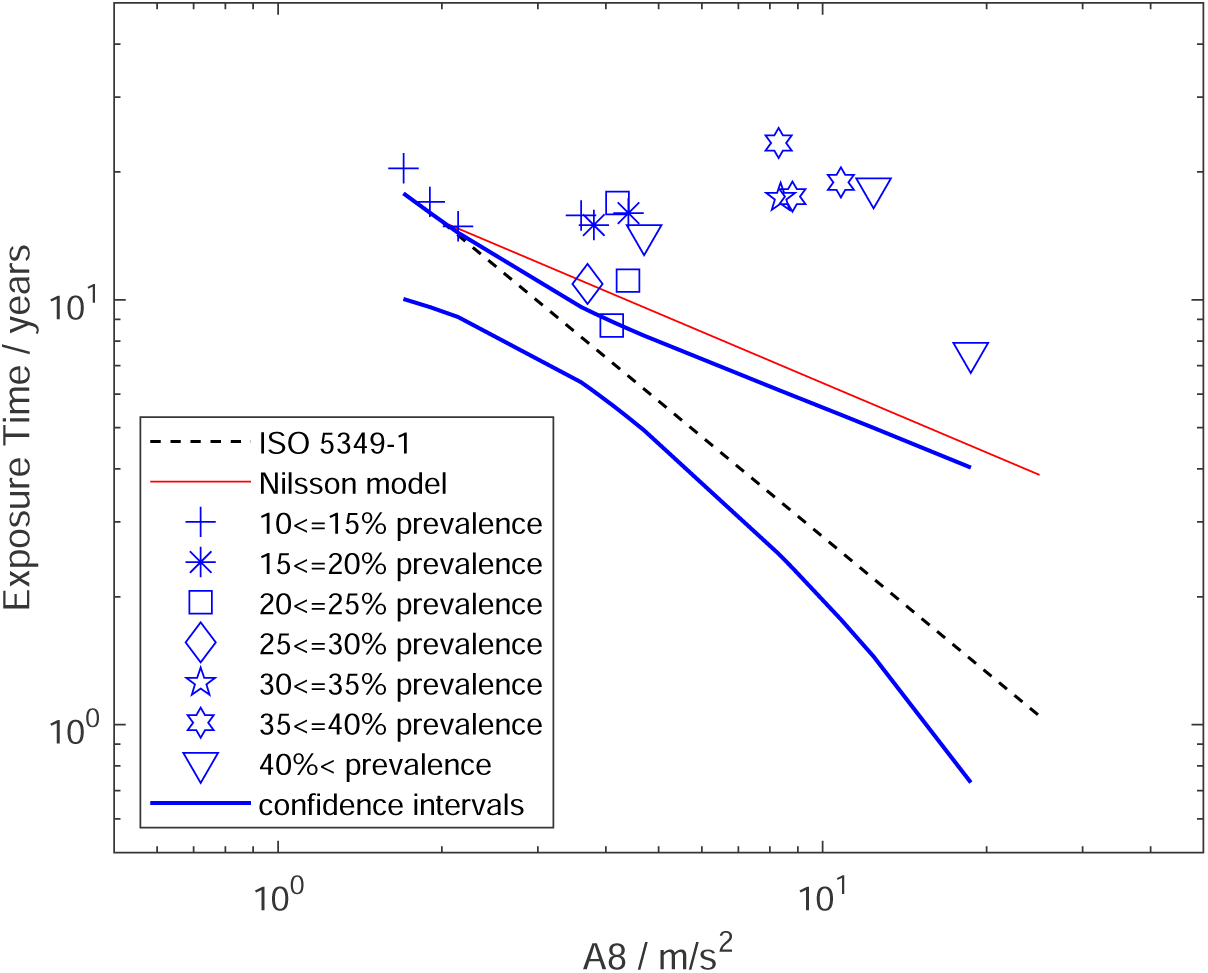
Mean exposure times versus A(8) reported in studies included in model 2 stratified by 5% prevalence intervals (see legend for symbols), all data points above exposure-response model from ISO 5349-1:2001 (dashed black line) and above or intersect the upper limit of the 95-percentile confidence interval of model 2 (thick blue lines), but not above model from Nilsson et al. (2017) (red line); note some data points with different prevalences cluster

Table 4 contains details of each model and the parameters of the regression analyses. The table identifies the data sources for each model and how the data were processed. The data from each study were given equal weight in all models. Values of *r*^2^ and parameters *a* and *b* of each regression analysis are given. In the studies included in the analyses shown in figures 2 and 4 workers used one or more of the following tools or machines: chipping hammers, straight grinders, rock drills, vertical grinders, hand cutters, rock breakers, angle grinders, light stone hammers, chainsaws, caulking tools, impact wrenches, nut runners, scaling hammers, hand-held grinders, polishers, road breakers, hammer drills, brush saws, and orbital sanders.

**Table 4.** Details of models including processing, *r*^2^-value for the regression analysis where applicable, fit parameters, and sources of the data included in each model (*b* for the model in ISO 5349:2001 is *−*1.07 (Brammer, 1982a))

|  |  |  |  |  |
| --- | --- | --- | --- | --- |
| Model | 1 | 2 | 3 | Original data |
| Figure | 1 | 2 | 3 | 4 |
| Raw prevalence adjusted to 10% | yes | yes | yes | no |
| Unedited data sorted by prevalence | no | no | no | yes |
| $r^2$ | 0.60 | 0.69 | - | - |
| Fit parameter $a$ | 16.5 | 20.6 | 10.4 | - |
| Fit parameter $b$ | $-0.57$ | $-0.74$ | $-0.63$ | - |
| Data sources | Bovenzi (1994), Bovenzi et al. (1995), Bovenzi (2008), Bovenzi et al. (2008), Bovenzi (2010a), Chatterjee et al. (1978) | Bovenzi (1994), Bovenzi et al. (1995), Bovenzi (1998b), Bovenzi (2008), Bovenzi et al. (2008), Bovenzi (2010a), Chatterjee et al. (1978) | Bovenzi et al. (1995), Bovenzi (2008), Chatterjee et al. (1978) | Bovenzi (1994), Bovenzi et al. (1995), Bovenzi (1998b), Bovenzi (2008), Bovenzi et al. (2008), Bovenzi (2010a), Chatterjee et al. (1978) |

Figure 1 shows the data set for which values of A(8) were provided by the authors of the studies, as well as the data point from Chatterjee et al. (1978) in which the A(8) value was derived from the component frequency spectra of the power tools. It can be seen from the figure that data are available for a broad range of frequency-weighted accelerations (from approximately A(8) = 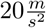), and lifetime exposures (from approximately 1.5 to 11 years). The data are scattered somewhat along the line of the model from the standard, a majority above and some below, while the line of the Nilsson et al. (2017) model lies above all data points. However, the regression line from model 1 runs roughly parallel to the latter and intersects the dashed line representing the ISO model at a value of A(8) of about 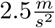. The 95 percentile lines, representing between them the most likely region in which the “true” relation between the lifetime exposure to reach 10% prevalence of VWF and A(8), include most data points and the ISO model (dashed black line). The red line representing the model from Nilsson et al. (2017) lies mostly outside the most probable region in which the “true” relation is expected to be found, and is intersected by the limit of one of the confidence intervals.

The results for model 2 are shown in figure 2. Here, the data from Bovenzi (1998b) for which additional calculations were needed are added to the data from the previous figure. The combined data set appears to be as scattered as that in figure 1. However, the slope of the regression line has shifted towards that of the model from the standard. From table 4 the gradients are now *−*0.74 and *−*1.07, respectively. As in figure 1, all the data points are below the red line that represents the model developed by Nilsson et al. (2017). The 95-percentile lines continue to enclose the ISO curve, but enclose fewer of the data points than previously. Thus the confidence intervals now define a much smaller region in which the “true” relation between exposure and the development of VWF is predicted to occur. This is reflected in the coefficient of determination, which has increased from 0.60 to 0.69 indicating that more of the variability has been captured by the model. No attempt has been made to further reduce the variability by introducing confounding variables or cofactors as none are considered in the ISO or Nilsson et al’s (2017) models. The red line representing the Nilsson et al. (2017) model now lies further outside the most probable region in which the “true” relation is expected, but is still intersected by one of the confidence intervals.

There are only three studies in which workers used only one power tool each workday in the meta-analysis of Nilsson et al. (2017) that can be included here. The data for these are shown in figure 3. With the low number of such studies, the possibilities for analysis are limited, but they can be fitted by a curve specified by equation 4. However, it lies far below the model in Nilsson et al. (2017), while the data points are closer to the model in the standard. Reference to table 4 reveals that the gradient for model 3 is close to that for model 1, namely *−*0.63 versus *−*0.57. However, there is a difference in *a*-values between model 3 and all the other models (viz., 10.4 versus 16.5 and 20.6).

This difference results in the regression line for model 3 falling partly outside the confidence intervals of model 2, which, from the data accepted here from the Nilsson et al. (2017)’s meta-analysis, define the most probable region in which the “true” relation between exposure and the development of 10% VWF prevalence is predicted to occur.

Finally, the original, unedited data from the studies used in the analyses are plotted in figure 4 according to the reported point prevalences, which have been divided into ranges (e.g., from 10 *to ≤* 15%, 15 *to ≤* 20%, etc.), and identified by different symbols. As already noted, all the studies included here have a prevalence of 10% or more. Hence, all the data points should be on or above a line that represents a model predicting 10% prevalence. This is the case for the dashed black line portraying the model from ISO 5349-1:2001, but not for the red line showing the model from Nilsson et al. (2017). The data can also be seen to lie above or intersect the upper 95-percentile confidence limit from model 2 (thick blue line), confirming that the confidence interval associated with this model does define a range of exposures within which VWF is expected to occur at a prevalence of 10% or less.

Close inspection of figure 4 reveals that data points with different prevalences cluster, implying unresolved issues remain in the method for calculating vibration exposure specified in the standard. The prevalences in these clusters range in one case from 10% to over 40% and in another from 25% to 40%. The single tool studies included in this study can be found as the triangle on the very right in figure 4 as well as the square just above the continuous red line representing the model from Nilsson et al. (2017) and the diamond on the latter. As their raw prevalences were 23.4% (square), 26.6% (diamond) and 50% (triangle), it would be expected that they would lie at increasing “distance” in time or A(8) from a line representing a 10% prevalence of VWF. This is at least roughly the case for the model from ISO 5349-1:2001, but less so for the model of Nilsson et al. (2017).

## 4 Discussion

A pooled analysis has been performed of studies identified as most likely to contain reliable data in a recent meta-analysis conducted by Nilsson et al. (2017). Additional selection rules have been introduced to control heterogeneity of the exposure data reported in different studies, which has been further reduced by re-calculating the lifetime exposure for studies in which a different metric was used from that employed here. Linear interpolation to a mean group prevalence of 10% has been used to reduce heterogeneity of the VWF point prevalences reported in different studies.

The PRISMA procedure Liberati et al (2009) was followed in the metaanalysis, with publications screened by abstract first. Out of the originally 4335 publications obtained by the literature search, 4041 were discarded. The authors did not give reasons for this high number of excluded studies. The remaining 294 publications were screened in full-text. Of these, 41 were deemed usable for an analysis of Raynaud’s phenomenon from exposure to vibration, as already noted, and another 11 studies were used to examine other health effects. The remaining 242 were excluded for reasons such as the aim of the study not being to evaluate the risk of HAVS, missing data on exposure or duplicate publishing of data. The chosen publications were then evaluated by a list of criteria designed to establish the risk of bias and hence the overall scientific “quality” of each study. The criteria weighted the subjective description of signs and symptoms, investigational methodology, differential diagnosis and staging of signs and symptoms. The weightings were not, however, incorporated in their derivation of the exposure-response model. For this the data from all studies included in the meta-analysis were deemed usable. By means of linear interpolation and extrapolation the mean exposure time at which there was a 10% prevalence of Raynaud’s phenomenon was determined. This was plotted versus the respective A(8)-value. Then a further analysis was performed to create a predictive model. The present analyses accepted the data sources believed by Nilsson et al. (2017) to contain low bias, and so avoided bias associated with the selection of studies by the present authors, but with some changes. Studies were screened using an additional set of selection rules introduced here to confirm the reliability of the data and compliance with the methods for evaluating vibration exposure in the international standards (table 1), and only those complying with both Nilsson et al.’s and these rules were included in the models. Furthermore, the calculation of the mean exposure time at 10% point prevalence was limited to interpolation, hence all studies with a raw prevalence of less than 10% were excluded from the models described here.

The reasons for the exclusion of extrapolated data from the analyses can be seen from figure 5. This diagram exemplifies the estimation of the mean lifetime exposures necessary for two notional population groups to reach 10% prevalence VWF, to illustrate the limitations of extrapolation for the type of models developed here. One notional population group had a prevalence of 25% VWF when the mean exposure of the group was 15 years, and the second a prevalence of 4% when the mean exposure was 8 years. The example requires interpolation for the former population group and extrapolation for the latter. The limitations of extrapolation may be illustrated by introducing uncertainty into the knowledge of the prevalence. In figure 5 the limitation takes the form of uncertainty concerning the magnitude of the observed, or raw, prevalence in each population group. This could arise, for example, from misdiagnosis, from subjects providing erroneous or misleading information (information bias), or from individuals being absent from the group at the time of a (cross-sectional) study. For the example in figure 5, the perturbation in the raw prevalence is taken to be *±*0.5% (i.e., an error involving one person in a population of 100 vibration-exposed individuals). The consequent uncertainty in the exposure durations estimated for 10% prevalence is shown by the thick horizontal blue line for interpolation and red line for extrapolation. Clearly, linear extrapolation to 10% prevalence from an observed prevalence below 10% introduces uncertainty of substantial magnitude into the estimated 10% prevalence compared to that introduced by linear interpolation.

**Fig. 5.**
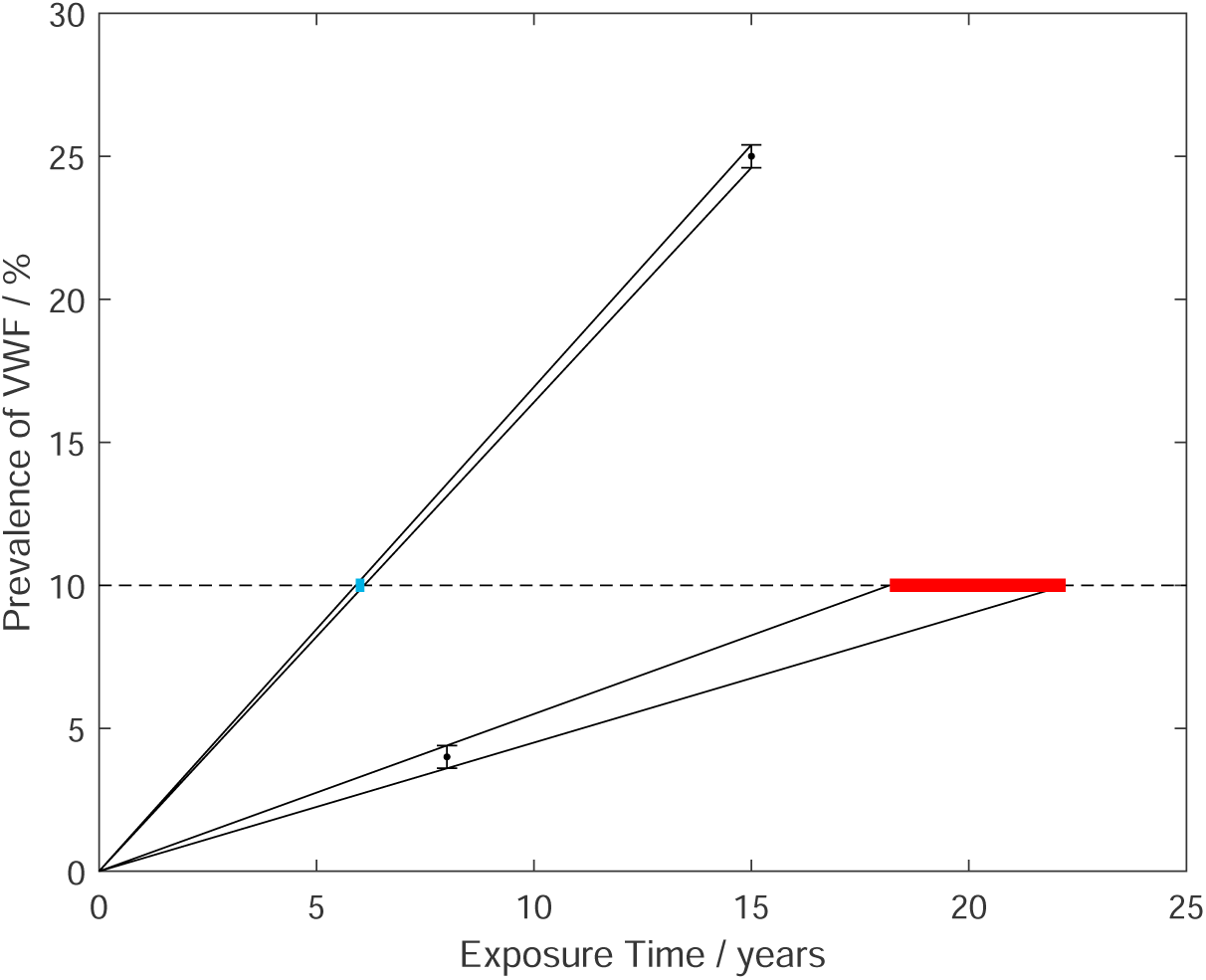
Two notional studies with equal uncertainties in prevalence (black circles with error bars), linear interpolation and extrapolation including uncertainties to 10% prevalence (black lines), effect of uncertainties on estimation of exposure time at which 10% prevalence occurs (blue interpolation, red extrapolation)

Comparing figures 1 and 2 to the analysis by Nilsson et al. (2017) reveals that using only interpolation eliminated data points from the models with mean lifetime exposures of 50 years, and more (see their figure 17). Such mean group lifetime exposure durations are highly unlikely for any occupation, and can only be obtained by some form of extrapolation. Hence, for the reasons described, extrapolation can be expected to introduce errors in the models with the inclusion of every data point so obtained. Even excluding studies with prevalence below 10%, the data are scattered (see, for example, figure 2). The model from Nilsson et al. (2017) is located above all data points and is not generally included within the 95-percentile confidence intervals for models 1 and 2. Hence it is not considered a probable fit to these data. This is believed to result primarily from the inclusion of extrapolated data in Nilsson et al.’s model. In contrast, the model from the international standard lies within the 95-percentile in both figures 1 and 2 and, as more data are included in the analysis, the closer the regression line approaches that in ISO 5349-1:2001. While the confidence intervals alone cannot confirm the validity of the model employed in ISO 5349-1:2001, neither can they confirm the need for its revision. Figure 4 demonstrates that the ISO model provides a conservative prediction for the development of 10% prevalence of VWF in a population group, as all data points lie above the line representing the model. This implies more exposure than depicted by the model in the ISO standard is needed to reach VWF prevalences of 10% or more. Furthermore, the 95-percentile confidence interval lies below or intersects all data points and does not generally include the model from Nilsson et al. (2017), yet encloses the model from ISO 5349-1:2001. As the interval identifies the region within which the exposure-response relation most probably lies and where the prevalence is expected to be 10% or less, it supports the conclusion that the prediction of the model from the standard is conservative. Yet, if the methods for evaluating vibration exposures contained in ISO 5349-1:2001 were generally applicable to all power tools and machines, and working conditions, the distribution of the data in figure 4 should be such that prevalence increases with increasing A(8) or lifetime exposure. However, data points with different prevalences and similar values of A(8) cluster, as already observed, implying that at least one parameter or factor in the construction of the vibration exposure (e.g., equation 1) needs to be revised.

When reliable data are selected for assessing the risk of developing VWF, figure 4 demonstrates that the ISO model provides a conservative prediction for the occurrence of 10% prevalence of VWF in a population group, as all data points lie above the line representing the model. This implies more exposure than depicted by the model in the ISO standard is needed to reach VWF prevalences of 10% or more. Furthermore, the 95-percentile confidence interval for 10% prevalence lies below or intersects all data points in figure 4 and does not generally include the model from Nilsson et al. (2017), yet encloses the model from ISO 5349-1:2001. As the interval identifies the region within which the exposure-response relation most probably lies and where the prevalence is expected to be 10% or, from data deemed reliable, less, it supports the conclusion that the prediction of the model from the standard is conservative. Yet, if the methods for evaluating vibration exposures contained in ISO 5349-1:2001 were generally applicable to all power tools and machines, and working conditions, the distribution of the data in figure 4 should be such that prevalence increases with increasing A(8) or lifetime exposure. However, data points with different prevalences and similar values of A(8) cluster, as already observed, implying that at least one parameter in the construction of the vibration exposure (e.g., equation 1) needs to be revised or an additional factor or confounder considered.

In considering the need for revision, it is important to distinguish between the exposure-response relation in ISO 5349-1:2001 and the methods for estimating the daily exposure. The former came from the model developed by Brammer (1982a,b), which considered epidemiologic studies involving workers whose full-time occupations involved near-daily, day-long operation of a single power tool or machine. No adjustment was made for the different daily duration of exposure occurring in different occupations. Methods for quantifying the daily exposure to hand-transmitted vibration were formulated independently from the model by the architects of ISO 5349-1:2001, and have not been modified in the analyses reported here. These include: 1) an equinoxious contour for exposure to vibration at different frequencies (i.e., a frequency weighting, included also in Brammer (1982a,b)); 2) the relative importance of the frequency-weighted acceleration and the daily duration of exposure in constructing the daily exposure; 3) the combination of exposures to different power tools or machines during a workday; and 4) the combination of daily exposures to construct a lifetime exposure. None of these factors has yet been considered in the models developed here, and each may have an effect on the resulting prediction.

The analyses reported here do provide insight into one of the factors influencing the quantification of exposure, namely the combining of exposures to different power tools or machines during a workday. According to the standard, when multiple tools are used during a working day, the measured exposure to each can be summed to obtain an overall daily exposure, expressed by the A(8)-value, according to equation 1. The ISO model is based on epidemiologic studies involving use of only one power tool or machine per day, as already noted, while the meta-analysis by Nilsson et al. (2017) contains only three such studies. In consequence, it is not possible to develop statistical inferences from these data. Nevertheless, it does appear by comparing the regression lines in figures 2 and 3 (or value for model 2 with those for model 3 in table 4) that the exposure-response relation for daily exposures using only single power tools or machines may deviate from that for daily exposures involving multiple power tools or machines constructed using equation 1. This implies the need to reconsider the calculation of daily exposures when multiple power tools or machines are used during a workday.

While the clustering of data in figure 4 also suggests that the method for combining exposures during a workday in equation 1 needs to be reconsidered, the scatter of data points when all prevalences have been adjusted to the same value suggests that broader reconsideration of the method for calculating exposure may be necessary (see figures 1 and 2). Inspection of the values for the coefficient of determination in table 4 reveals that the inclusion of additional epidemiologic studies (i.e., model 1 *→* 2) increased *r*^2^ from 0.60 to 0.69. The welcome, though modest, improvement in fit to the regression line is far short of that obtained in an analysis involving only exposures to single power tools during a workday (*r*^2^ = 0.82) (Brammer, 1982a). This last-mentioned analysis employed selection rules similar to those in table 1, but only included studies in which the prevalence of VWF was 50%, or greater.

The origin of the data scatter in figures 1 and 2 cannot be deduced from the results of the analyses presented here, but suggests that any re-appraisal of the calculation of daily exposure will require reconsideration of at least the other primary factor in its specification, namely the vibration magnitude(s). The apparent limitations of the frequency weighting employed in the international standard for assessing the harmful effects of hand-transmitted vibration have been well documented (Bovenzi (2012), Brammer and Pitts (2012), Griffin et al. (2003)). ISO has recently published a Technical Report that contains a frequency weighting specifically for assessing the risk of developing VWF, based on the analysis of Brammer and Pitts (2012) (ISO/TR 18570, 2017). Also, the time history of exposures, and in particular the impulsiveness that characterizes the vibration of impact tools, will need to be considered in future evaluations of exposure-response relations (Starck and Pyykkö (1986), Starck et al. (1990)).

The models derived here, as well as the model described by Nilsson et al. (2017), suffer from several limitations. Perhaps the most consequential is the estimation of the group mean lifetime exposure to reach 10% prevalence. The prevalence of VWF in a population group as exposure proceeds can be expected to follow a probability distribution dependent primarily on factors defining the health hazard to individuals and the number of persons in the group, combined with the changes in group membership. As the case definition of VWF is binary in nature, the period prevalence could be expected to approximately follow a cumulative normal distribution in a cohort with no change in membership, provided that there are a sufficient number of persons in the population group (ensured here by selection rule #6, table 1). Deviations from the anticipated distribution will result from persons entering and leaving the population group as exposure continues as well as changes in the daily exposure (e.g., from changes in work practices and in power tools or machines), with the magnitude of the deviations depending on these factors. However, the essential curvilinear form of the relation between point prevalence and exposure time can be expected to be maintained. Hence, linear interpolation, as used here and by Nilsson et al. (2017), will likely underestimate the lifetime exposure to 10% prevalence, and may render fortuitous the inclusion of the ISO prediction within the 95-percentile confidence intervals for the models. Consequently, future analyses will have to consider other methods of interpolation.

Another consideration is the correct identification of Raynaud’s phenomenon due to vibration. Selection rules #7 to #9 (table 1) have been introduced here to provide a common framework for assessing the epidemiologic data considered to contain low bias by Nilsson et al. (2017). The unintentional inclusion of individuals with signs and symptoms from causes other than vibration exposure in the observed prevalencealso tends to underestimate the lifetime exposure to 10% prevalence by linear interpolation. An additional consideration is determining the usage times for each power tool or machine used during a workday, and hence compiling a reasonable estimate of the daily exposure for the population group from observation or workers’ recollections. Clearly, with more power tools and machines used daily, and with normal day-to-day variations in work, this task multiplies, and the uncertainty in the daily exposure will increase.

A further limitation of our study arises from all but one of the publications employed in the analyses being conducted by one research group in a single country (Italy). This outcome of the process developed for selecting studies to include in the models was fortuitous. The selection rules were finalized before their application to any study was considered. Nevertheless, our results are subject to possible author bias and limited geographical applicability.

Nilsson et al. (2017) rated each study included in their meta-analysis based on a numerical score to assess the risk of bias, Of the 41 studies deemed acceptable for consideration of the development of VWF, the studies conducted by Bovenzi and co-workers used to construct our models were ranked from 2nd to 17th, with an average ranking of 8th (most reliable data ranked #1). Thus, there is little doubt that the studies are of high quality, and so are unlikely to contain significant author bias of a nature to invalidate their inclusion in a pooled analysis.

Reports of environmental conditions that precipitate episodic finger blanching have focused on a wide range of cool or cold temperatures as, for example, experienced in the United Kingdom or the continental USA, with the trigger mechanism also involving central body temperature, metabolic rate, vascular tone and emotional state (Taylor et al. (1975b), Hamilton (1918)). That VWF is repeatedly reported in Italy with its moderate climate would suggest that vibration-induced vascular disturbances are to be expected in countries at similar or increased latitudes. According to Nilsson et al. (2017), VWF has been reliably documented to have occurred in Canada, Finland, Italy, Japan, Korea, Sweden, The Netherlands, The United Kingdom, and the USA (see their Table 1). The fact that the international standard places no geographical restriction on the application of its exposure-response relation is further evidence that the primary causative agent is believed to be vibration rather than environmental, ethnic, or life-style factors peculiar to a single country (ISO 5349-1, 2001). The question of the universality of the vascular response to vibration, rather than the response being specific to a given country, has also been considered (Brammer, 1978). It was reasoned that the introduction and adoption world-wide of one-man chainsaws with similar technology in the early 1950s (Lee and Acres, 2020), and hence vibration, should lead to similar latencies of VWF in population groups of forest workers if the primary causative agent were vibration. In fourteen studies of full-time chainsaw operators published between 1964 and 1971, the mean latency for finger blanching was found to be 3.6 ± 1.0 years (mean ± standard deviation, SD). The short latency, commonly recorded more than ten years after introduction of the saws, together with the small SD, are suggestive of a vascular response that differs little between the countries in which the studies were conducted, which included Australia, Czechoslovakia, England, New Zealand, Scotland, and Tasmania in addition to many of the countries listed above. Thus our analyses are likely to be broadly applicable. However, the apparent absence of vibration-induced vasospasms being observed in a tropical climate has already been noted (Futatsuka et al. (2005), Su et al. (2013)).

## 5 Conclusions

Regression analyses have shown that excluding data points obtained by extrapolation and from studies failing the selection rules developed here changed the model for developing a 10% prevalence of VWF in a vibration-exposed population group from that proposed by Nilsson et al. (2017). Furthermore, it has been shown that without these data points the models derived here are closer to the model in the international standard, ISO 5349-1:2001. Hence, while the analyses cannot confirm the validity of the exposure-response relation in ISO 5349-1:2001, neither can they confirm the need for its revision. However, the analyses do confirm that the exposure-response relation proposed by Nilsson et al. (2017) does not improve the prediction of the prevalence of VWF in vibration-exposed population groups.

The range of exposures within which VWF is predicted to occur at prevalences of 10% or less has been derived in the form of a 95-percentile confidence interval. The model proposed by Nilsson et al. (2017) generally falls outside this interval and hence cannot be considered a fit to the epidemiologic data. In contrast, the ISO model is found to fall within the confidence interval and, as more studies are included in the models constructed here, the best fit to the data tends toward the ISO model although it still differs considerably in gradient.

The results of this study also demonstrate that the ISO model provides a conservative prediction for the development of 10% prevalence of VWF in a population group. They also reveal that the present method for evaluating vibration exposure contained in the international standard needs revision. Specifically, the models imply the need to revise the calculation of daily exposure when multiple power tools or machines are used during a workday. In future studies, alternate formulations of the vibration magnitude as well as predictive models that better represent the cumulative development of the prevalence of VWF in a group of workers will also be needed. And, finally, the data set will need to be expanded beyond the studies deemed usable in the meta-analysis by Nilsson et al. (2017), by including those not found by the search engines they used and those published since their meta-analysis was performed and in languages other than English.

Thus, at this time, we do not recommend changes to either the calculation of exposure or the exposure-response relation in ISO 5349-1:2001 (ISO 5349-1, 2001) until further analyses have been completed.

## Data Availability

All data produced in the present work are contained in the manuscript

## Declarations

## Ethics approval

Not applicable.

## Consent to participate

Not applicable.

## Consent for publication

Not applicable.

## Availability of data and materials

Not applicable.

## Code availability

Not applicable.

## Authors’ contributions

AB and MS conceived the study, MS did the data acquisition and statistical analysis. MS and AB drafted the manuscript. SM supervised and contributed to finalizing the manuscript.

## Acknowledgements

Not applicable.

